# The Lifetime Risk of Maternal Near Miss morbidity in Asia, Africa, the Middle East, and Latin America: a cross-country systematic analysis

**DOI:** 10.1101/2024.03.26.24304883

**Authors:** Ursula Gazeley, Antonino Polizzi, Julio Romero Prieto, José Manuel Aburto, Georges Reniers, Veronique Filippi

**Author notes:** Corresponding author, London School of Hygiene and Tropical Medicine, Keppel St, London WC1E 7HT.

## Abstract

**Background:** Life-threatening maternal near miss (MNM) morbidity can have long-term consequences for women’s physical, psychological, sexual, social, and economic wellbeing. The lifetime risk of MNM (LTR-MNM) quantifies the probability that a 15-year-old girl will experience a near miss before age 50, given current mortality and fertility levels. We compare LTR-MNM globally to reveal inequities in the cumulative burden of severe maternal morbidity across the reproductive life course.

**Methods:** We estimate the LTR-MNM for 40 countries with multi-facility, regional, or national data on the prevalence of MNM morbidity measured using World Health Organization (WHO) or modified WHO criteria of organ dysfunction from 2010 onwards (Central and Southern Asia=6, Eastern and South-Eastern Asia=9, Latin America and the Caribbean=10, Northern Africa and Western Asia=2, Sub-Saharan Africa=13). We also calculate the lifetime risk of severe maternal outcome (LTR-SMO) as the lifetime risk of maternal death or MNM.

**Findings:** The LTR-MNM ranges from a 1 in 1436 risk in China (2014) to 1 in 6 in Guatemala (2016), with a corresponding LTR-SMO from 1 in 887 to 1 in 5, respectively. The LTR-MNM is a 1 in 20 risk or higher in nine countries, seven of which are in sub-Saharan Africa. The LTR-SMO is a 1 in 20 risk or higher in 11 countries, eight of which are in sub-Saharan Africa. The relative contribution of the LTR-MNM to the LTR-SMO ranges from 42% in Angola to 99% in Japan.

**Interpretation:** There exists substantial global and regional inequity in the cumulative burden of severe maternal morbidity across the reproductive life course. The LTR-MNM is an important indicator to advocate for further global commitment to end preventable maternal morbidity. Finally, the LTR-SMO is an important tool to compare heterogeneity in the relative contribution of morbidity to the overall burden of maternal ill-health across the female reproductive life course, depending on countries’ stage in the obstetric transition.

**Funding:** This work was supported by U.G.’s PhD studentship from the UK Economic and Social Research Council [ES/P000592/1]. This work was also supported by the European Union Horizon 2020 research and innovation programme Marie Curie Fellowship (to J.M.A.) [grant agreement no. 896821], and Leverhulme Trust Large Centre Grant (to J.M.A. and A.P.).

**Research in Context:** *Evidence before this study:* We searched Embase, MEDLINE, and Global Health for English language studies reporting national, regional, or multi-facility estimates of the prevalence of life-threatening maternal morbidity (i.e., “maternal near miss” events), published from 2010 until 21 November 2023. Search terms included (1) “maternal near miss”/”severe (acute) maternal morbidity”/”life-threatening condition/complications” and (2) “prevalence”/”incidence”/ “ratio”/ “surveillance”. Our search revealed a dearth of population-level estimates: most existing prevalence data derive from (single) facility-based studies without accounting for births that occur outside of the facility. This bias may be substantial where institutional delivery rates are low. Second, existing global comparisons of the maternal near miss ratio indicate differences in the level of obstetric risk associated with an individual pregnancy only. But since women are at risk of experiencing a life-threatening complication with each pregnancy, existing data fail to account for differences in cumulative risk from repeat pregnancy. The lifetime risk of maternal near miss is a new indicator that attempts to address these deficits in the existing evidence base to better understand global inequities in the burden maternal near miss morbidity across women’s reproductive lives.

*Added value of this study:* We provide the first cross-country estimates of the lifetime risk of maternal near miss for 40 countries with multi-facility, regional, or national data on the prevalence of maternal near miss. We also calculate how the lifetime risk of maternal near miss compares to the lifetime risk of maternal death for a given country-year, and the relative contribution of morbidity to the lifetime risk of severe maternal outcome (the risk of death or near miss morbidity). This is the first study to do so.

*Implications of all the available evidence:* First, there is substantial global inequity in the risk of severe maternal morbidity across women’s reproductive lifetimes. By accounting for the cumulative risk from repeat pregnancy and women’s reproductive age survival, the lifetime risk of maternal near miss presents a clearer picture of cross-country disparities in the burden of near miss morbidity than prevalence data alone might suggest. Second, the composite risk that a girl will either die from a maternal cause or experience near miss morbidity during her lifetime is extremely high in many countries, particularly in sub-Saharan Africa. These findings provide a new lens through which to understand reproductive injustice, and a new opportunity to advocate for increased global commitment to end preventable maternal morbidity and mortality.

## Introduction

A maternal complication so severe that the woman almost died is called a maternal near miss (MNM). The World Health Organization (WHO) identifies MNM cases based on clinical, laboratory, and management-based indicators of organ dysfunction.^1^ These criteria are not, however, used universally and some countries use complication- or management-based criteria instead.^2^ Sharing many characteristics with the review of women who die from maternal causes, clinical audit of women who survive life-threatening complications is an effective tool to improve quality of maternal health care.^3^ Maternal near miss events reflect the ability of a healthcare system to save a woman’s life when life-threatening complications arise, and are testament to the importance of expanding access to and the quality of emergency obstetric care. However, surviving a complication of this severity can also lead to long-term physical, psycho-social, and economic sequelae.^4–7^ As countries progress through the obstetric transition ^8,9^, from high to low maternal mortality and direct obstetric to indirect (infectious and non-communicable) causes of maternal death, a greater proportion of adverse maternal outcomes are cases of near miss morbidity.

In response to global calls for better measurement of maternal morbidity,^10,11^ Gazeley et al (2023) proposed a new summary measure to compare the global burden of life-threatening maternal morbidity across women’s reproductive lifetimes.^12^ This new metric is called the “lifetime risk of maternal near miss” (LTR-MNM) and estimates the risk that a 15-year-old girl will experience a maternal near miss complication before age 50.^12^ Its measurement is analogous to the lifetime risk of maternal death (LTR-MD) – a metric used to compare countries and changes over time.^13^ Its intuitive appeal means the LTR-MNM fills an important gap in the measurement of maternal morbidity. Unlike the maternal near miss rate or ratio, the LTR-MNM estimates the cumulative risk of near miss morbidity. This is a function both the level of obstetric risk associated with an individual pregnancy (i.e., the maternal near miss ratio) and the risk from repeated exposure with each pregnancy (i.e., fertility levels), and all-cause mortality (to experience a near miss one must not die from a maternal cause or something else) ^12^.

When two lifetime risks – of death or near miss morbidity – are combined, the “lifetime risk of severe maternal outcome” (LTR-SMO) denotes the probability that a 15-year-old girl will either die from a maternal cause or experience a near miss event during her reproductive lifetime. This measure is an important tool to inform and strengthen global efforts to reduce all forms of preventable maternal mortality and morbidity.^12,14^

New research is needed to compare the LTR-MNM across countries to better understand global inequities in the burden of MNM morbidity over women’s reproductive lives. Our objective is to compare the global distribution of the LTR-MNM, the LTR-SMO, and heterogeneity in the relative contribution of morbidity vs. mortality to the LTR-SMO. This study is the first of its kind to do so.

## Methods

We used the GATHER statement to guide the reporting of our methods.^15^ All procedures were conducted using RStudio and are fully reproducible from open data. Our code is available at: https://github.com/polizzan/LTR-MNM-compare

### Calculation of the lifetime risk of maternal near miss

We calculated the LTR-MNM using the maternal near miss ratio for all reproductive ages 15-49 combined, following the procedure described in Gazeley et al. (2023). where age-disaggregated data on the maternal near miss ratio are not available.^12^ The formula for calculation is shown in Equation 1, where the [inlne] is the maternal near miss ratio for all ages 15-49 combined, NRR is the net reproduction rate, SRB is the sex ratio at birth, *l*_0_ is the initial female population radix (100,000), and *l*_15_ is the number of female survivors to age 15. For further detail on calculation, see Gazeley et al. (2023) ^12^.

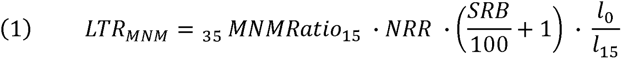

Second, we estimate the LTR-SMO as the summation of the lifetime risks of maternal death or near miss morbidity as follows:

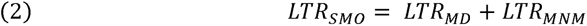

### Data inputs to Equations 1 and 2

#### 1. Maternal near miss data

##### a. Systematic search for MNM prevalence estimates

To derive national-level estimates of the LTR-MNM, we required data on the MNM ratio. Since our objective was to derive population-level estimates, and as the fertility and mortality data used to calculate the LTR-MNM are at the national level, we included only multi-facility, regional, or nationally representative data on the MNM ratio, excluding prevalence estimates derived from a single facility only. We also restricted eligible studies to those which identified MNM cases using either the full World Health Organization standard criteria of organ dysfunction,^1^ or a modified version of the WHO criteria for low-income settings (see section 1b below) to reduce noise in the MNM ratio due to heterogeneity in MNM criteria.

To identify eligible studies, we implemented two search strategies. First, we searched Embase, MEDLINE, and Global Health for studies reporting the prevalence of maternal near miss, with keywords for ‘national’, ‘regional’, or ‘population based’. Results were restricted to English language publications with reference periods from 2010 onwards, so that the data used to generate the LTR-MNM are still relatively current. The full search strategy is available in the Supplementary Material (Table S1). This search yielded 1174 results, of which 707 remained once duplicates were removed, and 117 were eligible for full text review. To avoid duplication of effort, our second strategy was to search the records included in several recent, global systematic reviews and meta-analyses for multi-facility, regional, or national studies of MNM prevalence. ^2,16–19^ In total, from these two search strategies we identified 41 studies (with 75 separate country estimates) eligible for inclusion. See Table S2 for the included studies.

##### b. Heterogeneity in the MNM criteria

There is little consistency in the criteria used to identify severe maternal morbidity cases.^2,20^ In 2009, the WHO developed a set of 25 clinical, laboratory, and management-based criteria of organ dysfunction in an attempt to standardise the measurement of MNM^1^. Where the laboratory or management capacity is lacking, the full WHO criteria can be hard to implement, and may miss true positive MNM. ^2,20–22^ Many studies therefore apply adaptations to the WHO organ dysfunction criteria to improve sensitivity in LMICs.^21–23^

Very few high-income countries use the WHO or alternative organ-dysfunction criteria, and instead often apply disease- and/or management-based criteria that are more readily available from administrative records without the need for additional data collection.^2,24^ With higher sensitivity but lower specificity, disease- and/or management based criteria typically result in higher estimates of the MNM ratio.^2,20,25^ These criteria may capture cases of morbidity that are less severe than the WHO criteria.

These differences in criteria introduce substantial heterogeneity in the measurement of the MNM ratio. To address this, we included only studies which applied either the WHO criteria or modified versions of the WHO organ dysfunction criteria in low-income settings. This aimed to ensure we are including estimates of the same severity of morbidity into the calculation of the lifetime risk, but also led to the exclusion of numerous studies from high-income countries. In instances where multiple organ dysfunction-based criteria were applied in the same study, we extracted and included each separate MNM estimate.

##### c. Denominator adjustment

The denominator of the MNM ratio is live births. For studies which used deliveries (n=4) or obstetric admissions (n=1) as the denominator, we calculated an approximate number of live births using (i) global data on the twin birth rate per 1000 deliveries from 2010-15 to partially account for multiple deliveries,^26^ and (ii) open access data on the stillbirth rate from UNICEF^27^. Second, most MNM ratio estimates derive from facilities. Since to survive a MNM most women require emergency intervention in a facility, a facility-level estimate of the number of MNM cases is likely a reasonable approximation of the true number of cases. However, for countries with low institutional delivery rates, facility-based estimates of live births in the MNM ratio denominator risk under-estimating the number of live births in a population. This potential bias is even greater if the MNM ratio derives only from tertiary referral facilities – which is the level of care women experiencing life-threatening morbidity require. To avoid over-estimating the MNM ratio, and consequently the LTR-MNM, we adjusted facility-based estimates of live births using open access data on the institutional delivery rate from the closest available year to studies’ reference period to account for births occurring outside of the facility (facility live births multiplied by the inverse of the institutional delivery rate).^12^

##### d. National-level estimation

To derive national estimates of the LTR-MNM, using fertility and mortality data, we first required a national estimate of the MNM ratio for each country. Where multiple multi-facility, regional, or national studies exist for a country, we used a random-effects only meta-analyses model (R package ‘metafor’ ^28^), with studies weighted by their sample size, to derive a single national-level MNM ratio estimate for each country. A random-effects only model was used given the heterogeneity in study designs, study populations and MNM criteria.^19^

#### 2. Additional data inputs on fertility and mortality levels

We used open-access estimates of age-specific fertility rates, *_n_f_x_*, survivors to age 15, *l_15_,* the NRR, and SRB, from the 2022 United Nations World Population Prospects (13) to calculate the LTR-MNM for each country with eligible MNM ratio data. To estimate the LTR-MD (and consequently the LTR-SMO), we used the latest WHO and Joint United Nations estimates of the maternal mortality rate (MMR) ^13^ (available <u>here</u>), alongside survival and fertility data from the World Population Prospects for consistency with the LTR-MNM.

### Uncertainty

We estimated uncertainty in the LTR-MNM deriving from uncertainty in the pooled MNM ratio estimate, keeping constant other sources of uncertainty (i.e., from WPP fertility and mortality estimates). We computed the 95% confidence intervals of the MNM ratio and the corresponding upper and lower bounds of the LTR-MNM.

## Results

In total, we estimated the MNM ratio for 40 countries with multi-facility, regional, or national data. The full results of the meta-analysis, including the number of studies per country, are available in the Supplementary Material (**Table S3).** Our meta-analysis estimate was then used to calculate the LTR-MNM, LTR-MD, and LTR-SMO. **Table 1** (below) presents the results for these 40 counties by their Sustainable Development Goal (SDG) regional grouping.

**Table 1:** Global estimates of the lifetime risk of maternal near miss, maternal death, and severe maternal outcome.

| Country | Year<sup>a</sup> | Total fertility rate | Maternal near miss ratio<sup>b</sup> | Maternal mortality ratio<sup>c</sup> | LTR-MNM 1 in N | LTR-MD 1 in N<sup>d</sup> | LTR-SMO 1 in N | Contribution of LTR-MNM to LTR-SMO (%) |
| --- | --- | --- | --- | --- | --- | --- | --- | --- |
| **Central and Southern Asia** |  |  |  |  |  |  |  |  |
| Afghanistan | 2010 | 6.1 | 7.1 | 898.7 | 24 | 19 | 11 | 44.3 |
| India | 2014 | 2.3 | 6.7 | 134.9 | 66 | 326 | 55 | 83.3 |
| Iran | 2014 | 2.0 | 8.7 | 20.9 | 57 | 2,372 | 56 | 97.6 |
| Nepal | 2012 | 2.4 | 2.1 | 287.7 | 206 | 148 | 86 | 41.7 |
| Pakistan | 2013 | 4.1 | 14.8 | 206.1 | 17 | 120 | 15 | 87.8 |
| Sri Lanka | 2010 | 2.2 | 4.0 | 37.3 | 114 | 1,234 | 104 | 91.6 |
| **Eastern and South-Eastern Asia** |  |  |  |  |  |  |  |  |
| Cambodia | 2010 | 2.8 | 10.6 | 276.4 | 35 | 134 | 28 | 79.3 |
| China | 2015 | 1.7 | 0.4 | 26.0 | 1,436 | 2,321 | 887 | 61.8 |
| Japan | 2010 | 1.4 | 5.9 | 5.7 | 122 | 12,788 | 121 | 99.1 |
| Laos | 2020 | 2.5 | 9.8 | 126.1 | 41 | 316 | 36 | 88.6 |
| Malaysia | 2014 | 2.1 | 2.2 | 22.5 | 222 | 2,146 | 201 | 90.6 |
| Mongolia | 2010 | 2.5 | 8.2 | 65.5 | 49 | 616 | 45 | 92.6 |
| Philippines | 2010 | 3.3 | 1.7 | 105.0 | 186 | 295 | 114 | 61.4 |
| Thailand | 2010 | 1.6 | 5.7 | 35.3 | 112 | 1,811 | 106 | 94.2 |
| Vietnam | 2010 | 1.9 | 2.0 | 87.6 | 269 | 608 | 186 | 69.3 |
| **Latin America and the Caribbean** |  |  |  |  |  |  |  |  |
| Argentina | 2012 | 2.3 | 4.8 | 45.0 | 89 | 958 | 82 | 91.5 |
| Brazil | 2011 | 1.8 | 10.0 | 61.9 | 56 | 904 | 53 | 94.1 |
| Ecuador | 2010 | 2.6 | 2.6 | 76.2 | 150 | 507 | 116 | 77.2 |
| Guatemala | 2016 | 3.0 | 61.9 | 103.1 | 6 | 330 | 5 | 98.4 |
| Honduras | 2014 | 2.6 | 11.8 | 68.3 | 33 | 561 | 31 | 94.5 |
| Mexico | 2010 | 2.3 | 11.1 | 51.2 | 39 | 841 | 37 | 95.6 |
| Nicaragua | 2010 | 2.6 | 13.2 | 97.8 | 30 | 397 | 28 | 93.1 |
| Paraguay | 2010 | 2.7 | 2.1 | 100.5 | 174 | 369 | 118 | 67.9 |
| Peru | 2010 | 2.6 | 10.0 | 76.4 | 40 | 515 | 37 | 92.9 |
| Suriname | 2018 | 2.4 | 12.9 | 97.6 | 32 | 428 | 30 | 93.0 |
| **Northern Africa and Western Asia** |  |  |  |  |  |  |  |  |
| Iraq | 2010 | 4.4 | 3.9 | 114.9 | 60 | 200 | 46 | 77.0 |
| Lebanon | 2010 | 2.1 | 4.3 | 18.0 | 109 | 2,630 | 105 | 96.0 |
| **Sub-Saharan Africa** |  |  |  |  |  |  |  |  |
| Angola | 2010 | 6.2 | 2.6 | 367.3 | 65 | 46 | 27 | 41.5 |
| Democratic Republic of Congo | 2013 | 6.5 | 19.7 | 584.6 | 8 | 28 | 6 | 77.2 |
| Ethiopia | 2018 | 4.3 | 13.5 | 311.9 | 18 | 76 | 14 | 81.3 |
| Ghana | 2016 | 3.9 | 26.9 | 258.1 | 10 | 103 | 9 | 91.2 |
| Kenya | 2015 | 3.8 | 4.5 | 483.0 | 62 | 57 | 30 | 48.0 |
| Namibia | 2018 | 3.5 | 8.2 | 218.0 | 37 | 138 | 29 | 79.0 |
| Niger | 2010 | 7.5 | 5.5 | 593.9 | 26 | 24 | 12 | 47.9 |
| Nigeria | 2014 | 5.7 | 11.3 | 1,135.3 | 17 | 17 | 8 | 49.9 |

| Country | Year<sup>a</sup> | Total fertility rate | Maternal near miss ratio<sup>b</sup> | Maternal mortality ratio<sup>c</sup> | LTR-MNM 1 in N | LTR-MD 1 in N<sup>d</sup> | LTR-SMO 1 in N | Contribution of LTR-MNM to LTR-SMO (%) |
| --- | --- | --- | --- | --- | --- | --- | --- | --- |
| South Africa | 2014 | 2.4 | 6.1 | 141.2 | 70 | 303 | 57 | 81.3 |
| Tanzania | 2012 | 5.1 | 22.3 | 393.7 | 9 | 52 | 8 | 85.0 |
| Uganda | 2012 | 5.8 | 13.6 | 334.4 | 13 | 54 | 11 | 80.2 |
| Zambia | 2016 | 4.7 | 13.0 | 155.4 | 17 | 142 | 15 | 89.3 |
| Zimbabwe | 2016 | 3.8 | 9.3 | 399.8 | 30 | 69 | 21 | 69.9 |

<sup>a</sup> Year is the average mid-point of the studies included in meta-analysis for each country.
<sup>b</sup> Meta-analysis pooled MNM ratio using adjusted denominators for facility-based estimates. Full adjustment and meta-analysis results can be found in Table S3.
<sup>c</sup> WHO and UN Joint Agency estimates of the MMR, matched according to closest year, per 100,000 live births.
<sup>d</sup> Authors' calculation of LTR-MD using WHO and UN Joint Agency MMR estimate for the given country-year.

In Central and Southern Asia, the LTR-MNM ranges from 1 in 206 (Nepal in 2012) to 1 in 17 (Pakistan in 2016); in Eastern and South-Eastern Asia from 1 in 1436 (China in 2014) to 1 in 35 (Cambodia in 2010); in Latin America, from 1 in 174 (Paraguay in 2010) to 1 in 6 (Guatemala in 2016); in Northern Africa and Western Asia, from 1 in 109 (Lebanon in 2010) to 1 in 60 (Iraq in 2010); in Sub-Saharan Africa, from 1 in 70 (South Africa in 2014) to 1 in 8 (Democratic Republic of Congo in 2016). The LTR-MNM is almost 240 times higher in Guatemala (the highest risk) than in China (the lowest risk).

Global variation in the LTR-MD is even greater than for the LTR-MNM, and ranges from 1 in 12,778 (Japan in 2010) to 1 in 17 (Nigeria in 2012), representing over a 750-fold difference in risk. Variation in the LTR-SMO – of experiencing either a MNM event of dying from a maternal cause – is still substantial, but less than for either the LTR-MNM or the LTR-MD. However, 11 countries had a LTR-SMO of at least 1 in 20 risk or higher; eight of these countries are in Sub-Saharan Africa.

Figure 1 shows the global heterogeneity in the LTR-MNM relative to countries’ fertility levels (Total Fertility Rate (TFR) from World Population Prospects) according to three quantile classes for each indicator, i.e. high, medium, and low LTR-MNM (>1 in 32, 1 in 32-65, <1 in 65 lifetime risk) and high, medium, and low TFR (>3.77, 2.42-3.77, <2.42 births per woman). Although most countries with a high LTR-MNM have a high TFR (dark magenta, e.g., Democratic Republic of Congo) and vice versa (light violet, e.g. China), there are some countries with a high LTR-MNM despite low fertility (dark red, e.g., Nicaragua).

**Figure 1.**
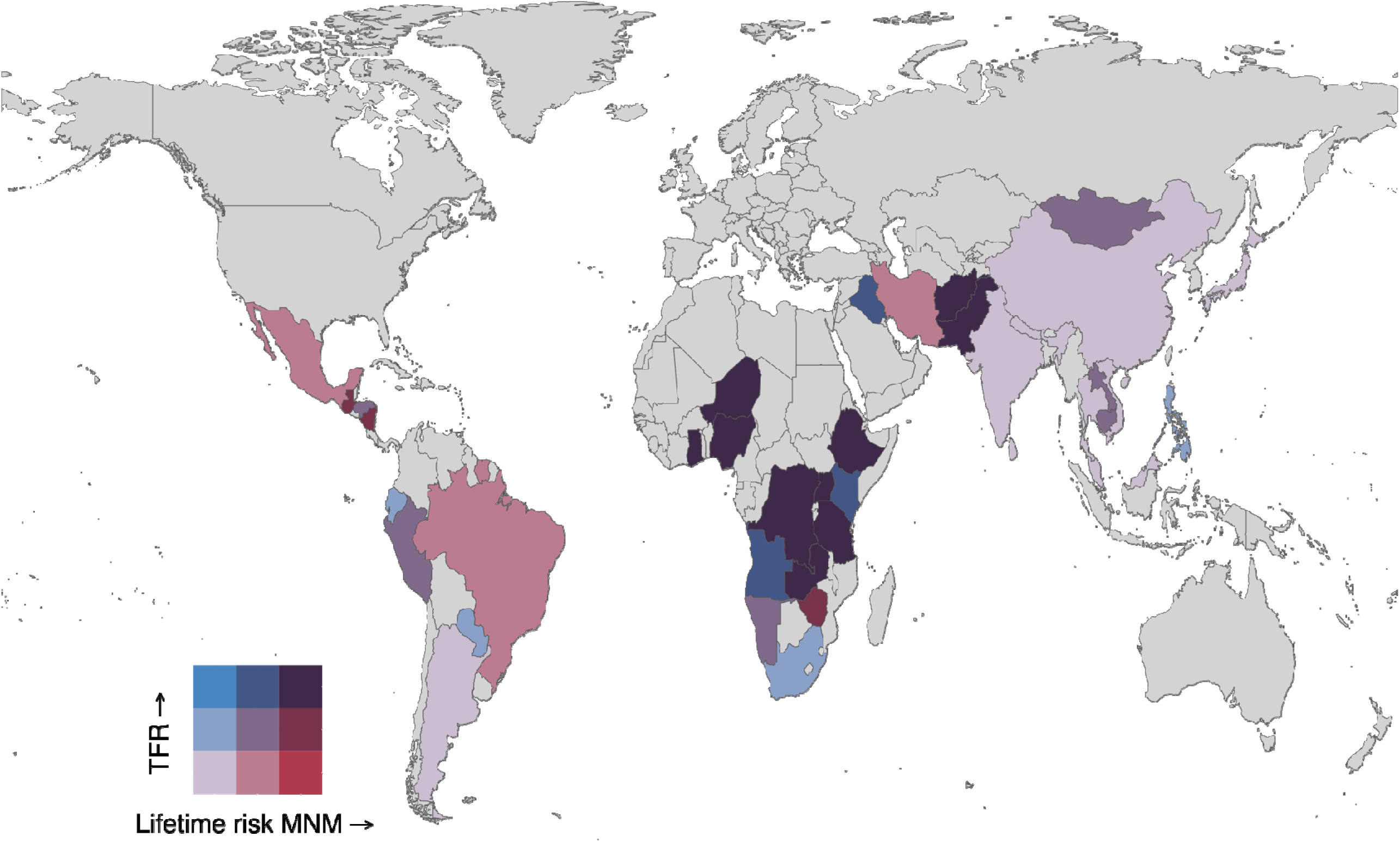
Global variation in the Lifetime Risk of Maternal Near Miss (LTR-MNM) and Total Fertility Rate (TFR)

Figure 2 shows the global inequity in the LTR-SMO and hence where the cumulative risk of experiencing either maternal death or maternal near miss morbidity is the highest. The burden of these two adverse maternal outcomes across women’s reproductive lifetimes is the highest among many countries in Sub-Saharan Africa, and some parts of Central and Southern Asia (Afghanistan and Pakistan, in particular).

**Figure 2.**
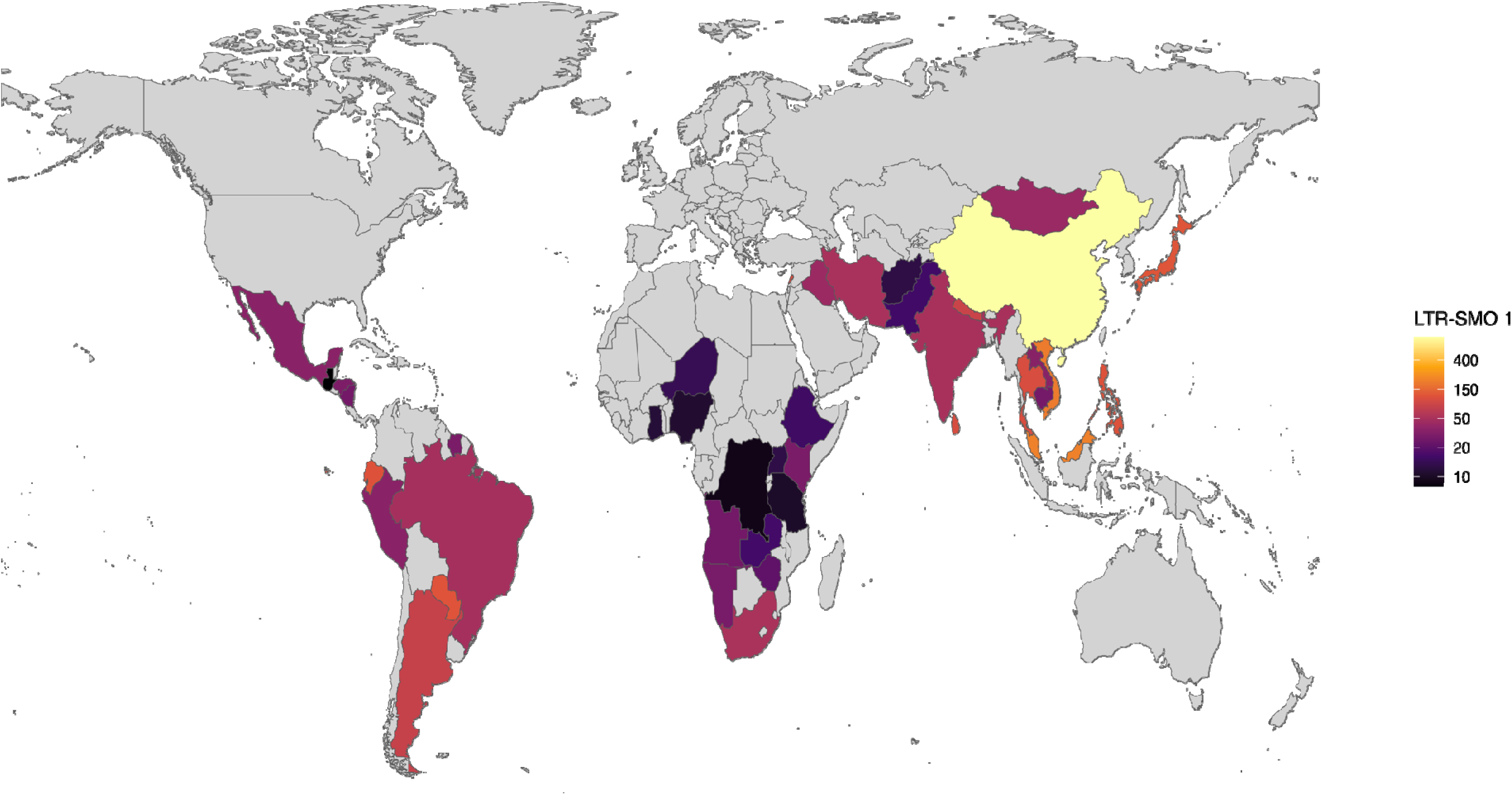
Global variation in the Lifetime Risk of Severe Maternal Outcome (LTR-SMO)

Variation in the contribution of LTR-MNM to the LTR-SMO according to countries’ positions in the obstetric transition is shown in Figure 3. For most countries in sub-Saharan Africa in Stage 1 or Stage 2 of the obstetric transition (MMR >500 or 300-499 per 100,000 live births, respectively), the contribution of near miss morbidity to the LTR-SMO is relatively low. However, as countries progress through the obstetric transition, the contribution of morbidity to the LTR-SMO is much greater. Some exceptions exist: China and Nepal have relatively low contributions of morbidity to the LTR-SMO for their stage in the obstetric transition.

**Figure 3.**
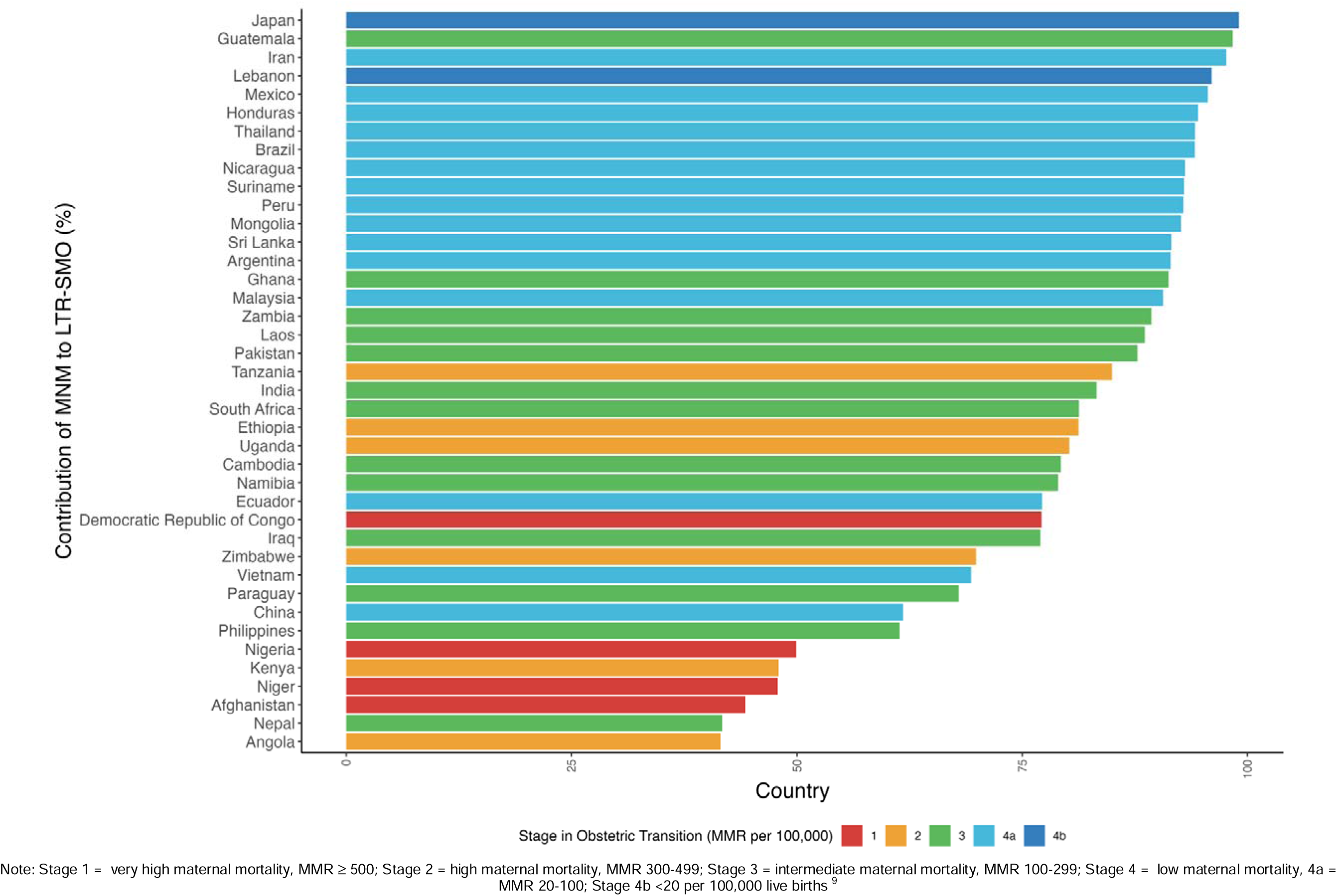
Contribution of near miss morbidity to Lifetime Risk of Severe Maternal Outcome by Stage in Obstetric Transition. Note: Stage 1 = very high maternal mortality, MMR ≥ 500; Stage 2 = high maternal mortality, MMR 300-499; Stage 3 = intermediate maternal mortality, MMR 100-299; Stage 4 = low maternal mortality, 4a = MMR 20-100; Stage 4b <20 per 100,000 live births ^9^

The relationship between countries’ LTR-MNM and their LTR-MD is available in the Supplementary Materials (Figure S1). On a log-log scale, there is a positive association between a countries’ LTR-MNM and their LTR-MD: countries with a high burden of maternal near miss morbidity are likely to also have a high burden of maternal mortality across the female reproductive life course.

### Sensitivity analysis

We calculated the LTR-MNM for estimates of the MNM without applying the denominator adjustment for facility-based studies. This adjustment makes a much greater difference in low resource contexts where the institutional delivery rate is low. In some countries, for example Ethiopia, the adjustment to the denominator for births occurring outside of facilities almost doubles the denominator, and therefore halves the MNM ratio (see Table S3). This downward adjustment of the level of obstetric risk therefore results in a lower estimate of the LTR-MNM than would if this adjustment was not applied. Full results of the sensitivity analysis are available in the Supplementary Material (Table S4).

### Uncertainty

Estimates of uncertainty in the LTR-MNM deriving from uncertainty in the MNM ratio is available in Table S5. Uncertainty in the LTR-MNM is substantial where there is a large degree of variability in the MNM ratio across studies.

## Discussion

We present the first ever cross-country estimates of the LTR-MNM – a new indicator that calculates the cumulative burden of severe maternal morbidity across the female reproductive life course. This measure addresses calls for more comparable measures of maternal morbidity. Unlike existing global comparisons of MNM prevalence, the LTR-MNM accounts for women’s repeated exposure to the risk of severe maternal morbidity with each pregnancy, and her survival throughout the reproductive ages 15-49. Further, capturing changes in the level of obstetric risk (MNM ratio), while accounting for prevailing fertility and mortality levels, means this is a better indicator of the burden of maternal morbidity in population.

Our results indicate that a 15-year-old girl in Guatemala has a 1 in 6 chance of experiencing a maternal near miss during her reproductive lifetime, and this is largely driven by a high (adjusted) MNM ratio estimate. A 15-year-old in the Democratic Republic of the Congo has a 1 in 8 chance, due to a moderate MNM ratio and high fertility levels. Finally, with an extremely low adjusted MNM ratio, and low fertility, we estimate that a 15-year-old girl in China has a 1 in 1436 chance of experiencing a near miss in her reproductive lifetime. This substantial inter- and intra-regional heterogeneity in the LTR-MNM highlights persistent inequities in maternal health outcomes. Global variation in the level of obstetric risk associated with an individual pregnancy (i.e. the MNM ratio) may reflect both low access to- and poor quality of- ante-, intra-, and post-partum care, and signify a health system’s capacity to identify and treat complications before they progress to become life-threatening.^2,3^ But the LTR-MNM also reveals how these inequities in obstetric risk are cumulative across the female reproductive Iife course. High fertility in many sub-Saharan African countries ^29^, and repeated exposure to severe maternal morbidity with each subsequent pregnancy, contributes to the high and extremely high LTR-MNM in many African countries, in particular. The LTR-MNM therefore presents a more accurate picture of the scale of global inequity in severe maternal morbidity than would be implied by differences in the MNM ratio alone.^12,14^

We also provide the first cross-country estimates of the LTR-SMO – the risk that a 15-year-old girl would experience either a maternal near miss complication or die from a maternity-related cause during her reproductive lifetime, once accounting for fertility and mortality levels in the population. This is an important tool for advocacy: most maternal near miss complications and almost all maternal deaths are preventable ^14^. The LTR-SMO emphasises the true burden of adverse maternal outcomes to women’s lives, their families, communities, and health systems, and the work still needed to end preventable forms of maternal morbidity and mortality ^12,14,30^.

The relative contribution of LTR-MNM to the LTR-SMO may be indicative of a country’s position in the obstetric transition – the secular shift from high to low maternal mortality, and direct to indirect causes of maternal death ^8,9^. As a country progresses through the obstetric transition, the capacity of their health care system to tackle severe complications when they arise and save women’s lives should improve with expansions in access to and the quality of emergency obstetric care. It may be expected, therefore, that the contribution of LTR-MNM to the LTR-SMO would be higher for countries which are further progressed through the obstetric transition. Figure 2 largely supports this. Outliers in this relationship may indicate possible near miss outperformers relative to their level of maternal mortality (e.g. Tanzania), as well as morbidity underperformers (e.g. Guatemala).

Finally, an unavoidable conclusion of our efforts to generate comparable estimates of the LTR-MNM is the urgent need for improved standardisation in the measurement of MNM globally.^2,20^ We estimate the LTR-MNM for all countries with national, regional, or multi-facility data on the MNM ratio if defined using (modified) WHO criteria of organ dysfunction. This decision was made to ensure we are measuring the same severity of maternal morbidity and reduce noise in the calculation of the LTR-MNM from MNM criteria. Many disease- and/or management-based criteria of severe maternal morbidity capture part of the morbidity spectrum closer to so-called “potentially life-threatening conditions”, that may or may not develop into life-threatening maternal near miss events. Studies using these broader criteria – predominantly from high income countries – were excluded to avoid substantial heterogeneity in MNM measurement biasing our LTR-MNM results.

The lack of standard MNM criteria implemented across all income settings means, therefore, that we are left with an incomplete picture of global inequities in the LTR-MNM, with Europe and North America unrepresented in our data. These are the countries where almost all the adverse outcomes are near miss events, and not maternal deaths, and hence where estimation of the LTR-MNM is imperative. Unlike most existing criteria used in high-income countries, the WHO near miss criteria do not use International Classification of Disease (ICD) codes, though ICD codes are routinely used in routine public health surveillance in most high income countries ^20^. As England et al. have argued, this likely contributes to the low uptake of the WHO criteria across high income settings. The application of ICD codes to the WHO criteria may facilitate measurement in countries’ routine administrative records or Health Management Information Systems. In turn, this may help to incentivise the uptake of the WHO criteria and improve the consistency of MNM measurement across income settings. Nevertheless, further efforts to promote the standardisation of MNM measurement are required to generate global estimates of the LTR-MNM across all income levels.

### Strengths and limitations

Although this study has multiple strengths – including its novelty, advancement of the evidence base for standard population-level indicators of maternal morbidity, and our attempts to standardise heterogeneous MNM measurement– it also has several limitations. First, some variation in the LTR-MNM within SDG regions are suggestive of residual heterogeneity in MNM criteria measurement that cannot easily be remedied, despite limiting the MNM estimates to those which used WHO-/modified WHO criteria of organ dysfunction only. Second, our approach to standardise study design differences (facility vs. population-level MNM ratio estimates) has a considerable effect on the estimated level of obstetric risk in some African populations. This heterogeneity in study design is typically not accounted for in meta-analyses of MNM. But in contexts with low institutional delivery rate, the potential underestimation of the denominator is substantial. This emphasises the need for more population-level data on severe maternal morbidity, especially in LMICs. Finally, although the LTR-MNM accounts for the cumulative effect of fertility, it does not account for potential non-linearity in this relationship (i.e., higher risk of MNM with higher parities) because parity specific MNM data are seldom available. This may therefore underestimate the true LTR-MNM in high fertility settings.

### Conclusion

Our findings expose substantial global and regional disparities in the burden of severe maternal morbidity across the female reproductive life span. These inequities are driven by differences in the level of obstetric risk, fertility levels, and reproductive age survival. The LTR-MNM and LTR-SMO are valuable indicators to emphasise the effect of severe adverse outcomes on women’s lives, communities, and health systems. These results underscore the need for the global community to redouble its efforts to end preventable maternal mortality and morbidity.

## Declarations

### Ethics approval

Ethics approval was not required to calculate the LTR-MNM as we used population-level data available in the public domain (published data on the MNM ratio and twin birth rate, and open-access fertility and mortality data from the United Nations World Population Prospects).

### Data availability

The full list of included MNM estimates are available in the supplementary material. Data on the twin birth rate are available from Monden et al (2021). All fertility and mortality data used in this article are freely available for download from the United Nations World Population Prospects Download Centre: https://population.un.org/wpp/Download/Standard/CSV/. All code is available at: https://github.com/polizzan/LTR-MNM-compare

### Author contributions

U.G. conceived the idea, developed the search strategy, ran the database searches, extracted the maternal near miss data, performed the computations, developed the code, developed the tables and visualisations, and drafted the initial manuscript. J.R.P., J.M.A., A.P., G.R. and V.F. supported the refinement of the study design, and the interpretation of results. A.P. developed the code and built the code repository. J.R.P., J.M.A., A.P., G.R. and V.F. revised the article.

## Acknowledgements

A.P. gratefully acknowledges the resources provided by the International Max Planck Research School for Population, Health and Data Science (IMPRS-PHDS).

