## Supplemental material for "The Lifetime Risk of Maternal Near Miss morbidity in Asia, Africa, the Middle East, and Latin America: a cross-country systematic analysis"

### A global comparison of the Lifetime Risk of Maternal Near Miss morbidity

#### Supplementary Material

#### Tables

#### Figures

Figure S1 Lifetime risk of maternal near miss versus lifetime risk of maternal death ........ 14

### 1. Search strategy

Table S1 includes the search terms used to identify eligible records in Embase, Global Health, and MEDLINE reporting multi-facility, regional, or national-level estimates of the prevalence of maternal near miss. This search was supplemented by also searching the included records of several recent meta-analyses on the prevalence of maternal near miss.

**Table S1 Search terms for eligible maternal near miss prevalence studies**

| Item number | Search term | Records retrieved |
|---|---|---|
| 1 | (prevalence or incidence or ratio or burden or surveillance).ti,ab. | 8558945 |
| 2 | ("population-based" or "region" or "regional" or "national").ti,ab. | 5257648 |
| 3 | ("maternal near miss maternal near-miss" or "severe acute maternal morbidity" or "SAMM" or "severe maternal morbidity" or "life-threatening complication" or "life-threatening maternal morbidity" or "life-threatening condition" or "life threatening complication" or "life threatening maternal morbidity" or "life threatening condition").ti,ab. | 40825 |
| 4 | 1 and 2 and 3 | 1315 |
| 5 | limit 4 to yr="2010 -Current" | 1202 |
| 6 | Limit 5 to English language | 1176 |
| 7 | Remove duplicates from 6 | 709<br>Embase: 661<br>Global Health: 27<br>MEDLINE: 21 |

#### 2. Maternal near miss prevalence studies included in meta-analysis

Table S2 (below) includes all studies included in our meta-analysis to estimate a single maternal near miss ratio (MNM ratio) per country with available multi-facility, regional, or national-level data. Studies were included only if (i) they used the World Health Organization (WHO) organ dysfunction criteria, or a modified version of the WHO criteria for low-income contexts; (ii) the reference period was from 2010 onwards. Where studies estimated the prevalence according to multiple WHO/modified organ dysfunction criteria, each estimate is included as a separate row of the meta-analyses.

**Table 2 Maternal near miss prevalence studies included in meta-analysis**

| Country | Author (Year) | Year of observation | Study design | Location | MNM criteria | MNM cases | Denominator | Type of denominator |
|---|---|---|---|---|---|---|---|---|
| **Central and Southern Asia** | | | | | | | | |
| Afghanistan | Souza et al. (2013) | 2010-11 | National | WHO Multicountry survey | WHO | 421 | 25,227 | livebirths |
| India | Goldenberg et al. (2017) | 2014-2016 | Regional | Surveillance area Belagavi - 18 primary health centers, 3 tertiary hospitals and 8 secondary hospitals Belagavi | Global Network | 615 | 21,548 | livebirths |
| India | Goldenberg et al. (2017) | 2014-2016 | Regional | Surveillance area Nagpur - 20 primary health centers, 10 tertiary hospitals and 129 secondary hospitals, Nagpur | Global Network | 79 | 17,541 | livebirths |
| India | Mansuri et al. (2019) | 2015-2016 | Regional | Four hospitals Ahmedabad | WHO | 247 | 21,491 | livebirths |
| India | Souza et al. (2013) | 2010-11 | National | WHO Multicountry survey | WHO | 174 | 30,094 | livebirths |
| Iran | Ghazivakili et al. (2016) | 2012 | Regional | All 13 public and private hospitals Alborz | WHO | 192 | 38,663 | livebirths |
| Iran | Hashemi et al. (2020) | 2016 | Regional | Five hospitals in Ahvaz | WHO | 81 | 3,002 | livebirths |
| Iran | Naderi et al. (2015) | 2013 | Regional | Eight hospitals in Southeast Iran | WHO | 501 | 19,908 | livebirths |
| Nepal | Rana et al. (2013) | 2012 | Regional | 9 facilities Kathmandu valley | WHO | 157 | 41,676 | livebirths |
| Nepal | Souza et al. (2013) | 2010-11 | National | WHO Multicountry survey | WHO | 65 | 10,999 | livebirths |
| Pakistan | Goldenberg et al. (2017) | 2014-2016 | Regional | Surveillance area - 47 primary health clinics, 25 secondary care facilities and 3 referral hospitals, Thatta district | Global Network | 1,830 | 21,604 | livebirths |
| Pakistan | Souza et al. (2013) | 2010-11 | National | WHO Multicountry survey | WHO | 94 | 12,729 | livebirths |

| Country | Author (Year) | Year of observation | Study design | Location | MNM criteria | MNM cases | Denominator | Type of denominator |
|---|---|---|---|---|---|---|---|---|
| Sri Lanka | Souza et al. (2013) | 2010-11 | National | WHO Multicountry survey | WHO | 73 | 17,988 | livebirths |
| **Eastern and South-Eastern Asia** | | | | | | | | |
| Cambodia | Souza et al. (2013) | 2010-11 | National | WHO Multicountry survey | WHO | 59 | 4,635 | livebirths |
| China | Carmen et al. (2023) | 2019 | Regional | Three tertiary centres in Hong Kong | WHO | 61 | 11,075 | livebirths |
| China | Li et al. (2022) | 2017-2018 | Regional | 18 hospitals southern china | WHO but unclear | 417 | 138,556 | pregnant women |
| China | Ma et al. (2020) | 2012-2017 | Regional | 18 hospitals in Zhejiang province | WHO | 3,208 | 543,109 | livebirths |
| China | Souza et al. (2013) | 2010-11 | National | WHO Multicountry survey | WHO | 34 | 13,242 | livebirths |
| China | Xiong et al. (2020) | 2012-2018 | Regional | 17 hospitals in Hunan province | WHO | 489 | 511,793 | livebirths |
| China | Yi Mu et al. (2019) | 2012-2017 | National | NA | WHO | 37,060 | 90,522,716 | livebirths |
| Japan | Souza et al. (2013) | 2010-11 | National | WHO Multicountry survey | WHO | 21 | 3,527 | livebirths |
| Laos | Luexay et al. (2014) | 2010 | Regional | 11 districts in Sayaboury province | Global Network | 11 | 1,122 | livebirths |
| Malaysia | Norhayati et al. (2016) | 2014 | Regional | 2 facilities in Kelantan | WHO | 47 | 21,579 | livebirths |
| Mongolia | Souza et al. (2013) | 2010-11 | National | WHO Multicountry survey | WHO | 61 | 7,303 | livebirths |
| Philippines | Souza et al. (2013) | 2010-11 | National | WHO Multicountry survey | WHO | 29 | 10,609 | livebirths |
| Thailand | Souza et al. (2013) | 2010-11 | National | WHO Multicountry survey | WHO | 51 | 8,894 | livebirths |
| Vietnam | Souza et al. (2013) | 2010-11 | National | WHO Multicountry survey | WHO | 33 | 15,411 | livebirths |
| **Latin America and the Caribbean** | | | | | | | | |
| Argentina | Abalos et al. (2014) | 2012 | Regional | NA | WHO | 28 | 6,024 | livebirths |
| Argentina | De Mucio et al. (2016) | 2013-2014 | Regional | 3 hospitals with >3000 deliveries a year | WHO | 2 | 762 | livebirths |
| Argentina | Souza et al. (2013) | 2010-11 | National | WHO Multicountry survey | WHO | 51 | 9,729 | livebirths |

| Country | Author (Year) | Year of observation | Study design | Location | MNM criteria | MNM cases | Denominator | Type of denominator |
|---|---|---|---|---|---|---|---|---|
| Brazil | Dias et al. (2014) | 2011-2012 | National | NA | WHO | 23,737 | 2,325,394 | livebirths |
| Brazil | Galvao et al. (2014) | 2011-2012 | Regional | The two only maternity hospitals for the entire state: Santa Izabel Hospital and Nossa Senhora de Lourdes Maternity. | WHO | 77 | 16,243 | livebirths |
| Brazil | Menezes et al. (2015) | 2011-2012 | Regional | 2 hospitals in Aracaju | WHO | 77 | 20,435 | admissions |
| Brazil | Souza et al. (2013) | 2010-11 | National | WHO Multicountry survey | WHO | 17 | 7,019 | livebirths |
| Ecuador | Souza et al. (2013) | 2010-11 | National | WHO Multicountry survey | WHO | 30 | 10,108 | livebirths |
| Guatemala | Goldenberg et al. (2017) | 2014-2016 | Regional | Surveillance area - 1 referral hospital, 30 health centers, and 42 health posts, Chimaltenango region | Global Network | 1,221 | 19,712 | livebirths |
| Honduras | De Mucio et al. (2016) | 2013 | Regional | 2 hospitals with >3000 annual deliveries | WHO | 10 | 613 | livebirths |
| Mexico | Souza et al. (2013) | 2010-11 | National | WHO Multicountry survey | WHO | 153 | 13,167 | livebirths |
| Nicaragua | Souza et al. (2013) | 2010-11 | National | WHO Multicountry survey | WHO | 119 | 6,426 | livebirths |
| Paraguay | Souza et al. (2013) | 2010-11 | National | WHO Multicountry survey | WHO | 8 | 3,595 | livebirths |
| Peru | Souza et al. (2013) | 2010-11 | National | WHO Multicountry survey | WHO | 169 | 15,021 | livebirths |
| Suriname | Verschueren et al. (2020) | 2017-2018 | National | NA | WHO | 71 | 9,114 | livebirths |
| Suriname | Verschueren et al. (2020) | 2017-2018 | National | NA | WHO modification for Namibia | 118 | 9,114 | livebirths |

| Country | Author (Year) | Year of observation | Study design | Location | MNM criteria | MNM cases | Denominator | Type of denominator |
|---|---|---|---|---|---|---|---|---|
| Suriname | Verschueren et al. (2020) | 2017-2018 | National | NA | WHO modification for Sub-Saharan Africa | 242 | 9,114 | livebirths |
| **Northern Africa and Western Asia** | | | | | | | | |
| Iraq | Jabir et al. (2012) | 2010 | Regional | 6 hospitals in Baghdad | WHO | 128 | 25,472 | livebirths |
| Lebanon | Souza et al. (2013) | 2010-11 | National | WHO Multicountry survey | WHO | 18 | 4,008 | livebirths |
| **Sub-Saharan Africa** | | | | | | | | |
| Angola | Souza et al. (2013) | 2010-11 | National | WHO Multicountry survey | WHO | 57 | 9,966 | livebirths |
| Democratic Republic of Congo | Goldenberg et al. (2017) | 2014-2016 | Regional | Surveillance area | Global Network | 521 | 13,637 | livebirths |
| Democratic Republic of Congo | Souza et al. (2013) | 2010-11 | National | WHO Multicountry survey | WHO | 88 | 8,395 | livebirths |
| Ethiopia | Beyene et al. (2022) | 2018 | Regional | Three hospitals in southern Ethiopia | WHO | 90 | 2,880 | livebirths |
| Ethiopia | Gebremariam et al. (2022) | 2012-2017 | Regional | Three selected hospitals in North Shewa Zone, Central Ethiopia | WHO | 129 | 905 | pregnant women |
| Ethiopia | Kusheta et al. (2023) | 2019 | Regional | All public hospitals in Hadiya zone, southern Ethiopia | sub-Saharan Africa criteria | 70 | 2,724 | livebirths |
| Ethiopia | Tenaw et al. (2021) | 2019-2020 | Regional | Two major private hospitals in Harar and Dire Dawa | WHO modification for Sub-Saharan Africa | 108 | 1,173 | livebirths |
| Ethiopia | Tura et al. (2018) | 2016-2017 | Regional | Two hospitals in Eastern Ethiopia | sub-Saharan Africa criteria | 594 | 7,404 | livebirths |
| Ethiopia | Tura et al. (2018) | 2016-2017 | Regional | Two hospitals in Eastern Ethiopia | WHO | 128 | 7,404 | livebirths |
| Ethiopia | Wakgar et al. (2019) | 2014-2016 | Regional | Hawassa University comprehensive specialized and Yirgalem hospital. | WHO | 501 | 15,059 | admissions |

| Country | Author (Year) | Year of observation | Study design | Location | MNM criteria | MNM cases | Denominator | Type of denominator |
|---|---|---|---|---|---|---|---|---|
| Ethiopia | Worke et al. (2019) | 2018 | Regional | Three out of five referral hospitals in Amhara chosen randomly | WHO | 152 | 572 | admissions |
| Ethiopia | Yemane et al. (2020) | 2017 | Regional | Three randomly selected public hospitals in south western Ethiopia | WHO | 210 | 5,530 | livebirths |
| Ghana | Oppong et al. (2018) | 2015 | Regional | Three tertiary hospitals in Southern Ghana | Modified WHO (Ghana) | 288 | 8,433 | livebirths |
| Kenya | Goldenberg et al. (2017) | 2014-2016 | Regional | Surveillance area | Global Network | 433 | 13,724 | livebirths |
| Kenya | Owolabi et al. (2020) | 2018 | National | NA | WHO | 5,116 | 708,459 | livebirths |
| Kenya | Souza et al. (2013) | 2010-11 | National | WHO Multicountry survey | WHO | 77 | 19,658 | livebirths |
| Namibia | Heemelaar et al. (2019) | 2018 | Regional | Four representative hospitals | WHO | 61 | 5,772 | livebirths |
| Namibia | Heemelaar et al. (2019) | 2018 | Regional | Four representative hospitals | Modified WHO (Namibia) | 184 | 5,772 | livebirths |
| Namibia | Heemelaar et al. (2020) | 2018-2019 | National | NA | Modified WHO (Namibia) | 298 | 37,106 | livebirths |
| Niger | Souza et al. (2013) | 2010-11 | National | WHO Multicountry survey | WHO | 196 | 10,714 | livebirths |
| Nigeria | Adanikin et al. (2019) | 2012-2013 | National | NA | WHO | 1,451 | 91,724 | livebirths |
| Nigeria | Souza et al. (2013) | 2010-11 | National | WHO Multicountry survey | WHO | 298 | 11,775 | livebirths |
| Nigeria | Tukur et al. (2022) | 2019-2020 | National | NA | WHO | 5,678 | 69,055 | livebirths |
| South Africa | Heitkamp et al. (2021) | 2014-2015 | Regional | Metro east cape town | WHO | 268 | 32,161 | livebirths |
| South Africa | Iwuh et al et al. (2018) | 2014 | Regional | Metro west cape town | WHO | 112 | 19,222 | livebirths |

| Country | Author (Year) | Year of observation | Study design | Location | MNM criteria | MNM cases | Denominator | Type of denominator |
|---|---|---|---|---|---|---|---|---|
| South Africa | Soma-pillay et al. (2017) | 2013-2014 | Regional | Tshwane SA | WHO | 117 | 26,614 | pregnant women |
| Tanzania | Litorp et al. (2014) | 2012 | Regional | Two hospitals, dar es Salaam | WHO | 467 | 13,121 | livebirths |
| Uganda | Nakimuli et al. (2016) | 2013-2014 | Regional | Two referral hospitals | WHO | 695 | 25,840 | livebirths |
| Uganda | Souza et al. (2013) | 2010-11 | National | WHO Multicountry survey | WHO | 120 | 10,467 | livebirths |
| Zambia | Goldenberg et al. (2017) | 2014-2016 | Regional | Surveillance area | Global Network | 167 | 12,827 | livebirths |
| Zimbabwe | Chikadaya et al. (2018) | 2016 | Regional | Two referral hospitals for all of Harare | WHO | 110 | 11,871 | livebirths |

#### 3. Meta-analysis results

We used a random effects only meta-analyses to calculate a single maternal near miss ratio for each country with available multi-facility, regional, or national-level data on the prevalence of near miss. The full results can be found in Table S3 below. Adjusted results present the pooled MNM ratio used in our calculation of the lifetime risk of maternal near miss (LTR-MNM) and lifetime risk of severe maternal outcome (LTR-SMO). For these estimates, we have adjusted the denominator for facility-based studies (see Table S2) using the institutional delivery rate, to better account for total births, including those occurring outside of facilities.

To highlight the effect of our adjustment, the unadjusted results present the meta-analysis pooled MNM ratio where facility-based studies have not been adjusted by the institutional delivery rate.

**Table 3 Meta-analysis results for estimation of the maternal near miss ratio**

| | ISO | Country | Year midpoint | No. of studies | Unadjusted MNM ratio | Adjusted MNM ratio |
|---|---|---|---|---|---|---|
| **Central and Southern Asia** | | | | | | |
| | AFG | Afghanistan | 2010 | 1 | 16.69 | 7.14 |
| | IND | India | 2014 | 4 | 9.48 | 6.71 |
| | IRN | Iran | 2014 | 3 | 9.11 | 8.68 |
| | NPL | Nepal | 2012 | 2 | 4.14 | 2.06 |
| | PAK | Pakistan | 2013 | 2 | 34.28 | 14.82 |
| | LKA | Sri Lanka | 2010 | 1 | 4.06 | 4.04 |
| Mean | — | — | — | — | 13.0 | 7.2 |
| **Eastern and South-Eastern Asia** | | | | | | |
| | KHM | Cambodia | 2010 | 1 | 12.73 | 10.59 |
| | CHN | China | 2015 | 6 | 0.42 | 0.42 |
| | JPN | Japan | 2010 | 1 | 5.95 | 5.94 |
| | LAO | Laos | 2020 | 1 | 9.80 | 9.80 |
| | MYS | Malaysia | 2014 | 1 | 2.18 | 2.18 |
| | MNG | Mongolia | 2010 | 1 | 8.35 | 8.23 |
| | PHL | Philippines | 2010 | 1 | 2.73 | 1.67 |
| | THA | Thailand | 2010 | 1 | 5.73 | 5.71 |
| | VNM | Vietnam | 2010 | 1 | 2.14 | 1.98 |
| Mean | — | — | — | — | 5.6 | 5.2 |
| **Latin America and the Caribbean** | | | | | | |
| | ARG | Argentina | 2014 | 3 | 4.86 | 4.83 |
| | BRA | Brazil | 2012 | 4 | 10.06 | 9.96 |
| | ECU | Ecuador | 2010 | 1 | 2.97 | 2.58 |
| | GTM | Guatemala | 2016 | 1 | 61.94 | 61.94 |
| | HND | Honduras | 2014 | 1 | 16.31 | 11.75 |
| | MEX | Mexico | 2010 | 1 | 11.62 | 11.11 |
| | NIC | Nicaragua | 2010 | 1 | 18.52 | 13.15 |
| | PRY | Paraguay | 2010 | 1 | 2.23 | 2.13 |
| | PER | Peru | 2010 | 1 | 11.25 | 9.97 |
| | SUR | Suriname | 2018 | 3 | 13.89 | 12.90 |
| Mean | — | — | — | — | 15.4 | 14.0 |
| **Northern Africa and Western Asia** | | | | | | |
| | IRQ | Iraq | 2010 | 1 | 5.03 | 3.85 |
| | LBN | Lebanon | 2010 | 1 | 4.49 | 4.34 |
| Mean | — | — | — | — | 4.8 | 4.1 |
| **Sub-Saharan Africa** | | | | | | |
| | AGO | Angola | 2010 | 1 | 5.72 | 2.61 |
| | COD | Democratic Republic of Congo | 2013 | 2 | 23.34 | 19.74 |
| | ETH | Ethiopia | 2018 | 8 | 42.81 | 13.52 |
| | GHA | Ghana | 2016 | 1 | 34.15 | 26.88 |
| | KEN | Kenya | 2015 | 3 | 7.30 | 4.45 |
| | NAM | Namibia | 2018 | 2 | 8.33 | 8.20 |

| | ISO | Country | Year midpoint | No. of studies | Unadjusted MNM ratio | Adjusted MNM ratio |
|---|---|---|---|---|---|---|
| | NER | Niger | 2010 | 1 | 18.29 | 5.45 |
| | NGA | Nigeria | 2014 | 3 | 31.59 | 11.31 |
| | ZAF | South Africa | 2014 | 3 | 6.14 | 6.14 |
| | TZA | Tanzania | 2012 | 1 | 35.59 | 22.28 |
| | UGA | Uganda | 2012 | 2 | 21.03 | 13.57 |
| | ZMB | Zambia | 2016 | 1 | 13.02 | 13.02 |
| | ZWE | Zimbabwe | 2016 | 1 | 9.27 | 9.27 |
| Mean | — | — | — | — | 19.7 | 12.0 |

#### 4. Lifetime risk of maternal near miss vs. death

Figure S1 plots the lifetime risk of maternal near miss against the lifetime risk of maternal death, on a log-log scale. A log-log scale was chosen due to the data spans several orders of magnitude between the lowest observed LTR-MNM or LTR-MD and the highest values. There is a positive association between these two indicators: the higher the LTR-MNM (smaller number), the higher the LTR-MD (smaller number). Some exceptions exist, indicating morbidity underperformers for their LTR-MD (e.g. Guatemala).

#### Figure S1 Lifetime risk of maternal near miss versus lifetime risk of maternal death

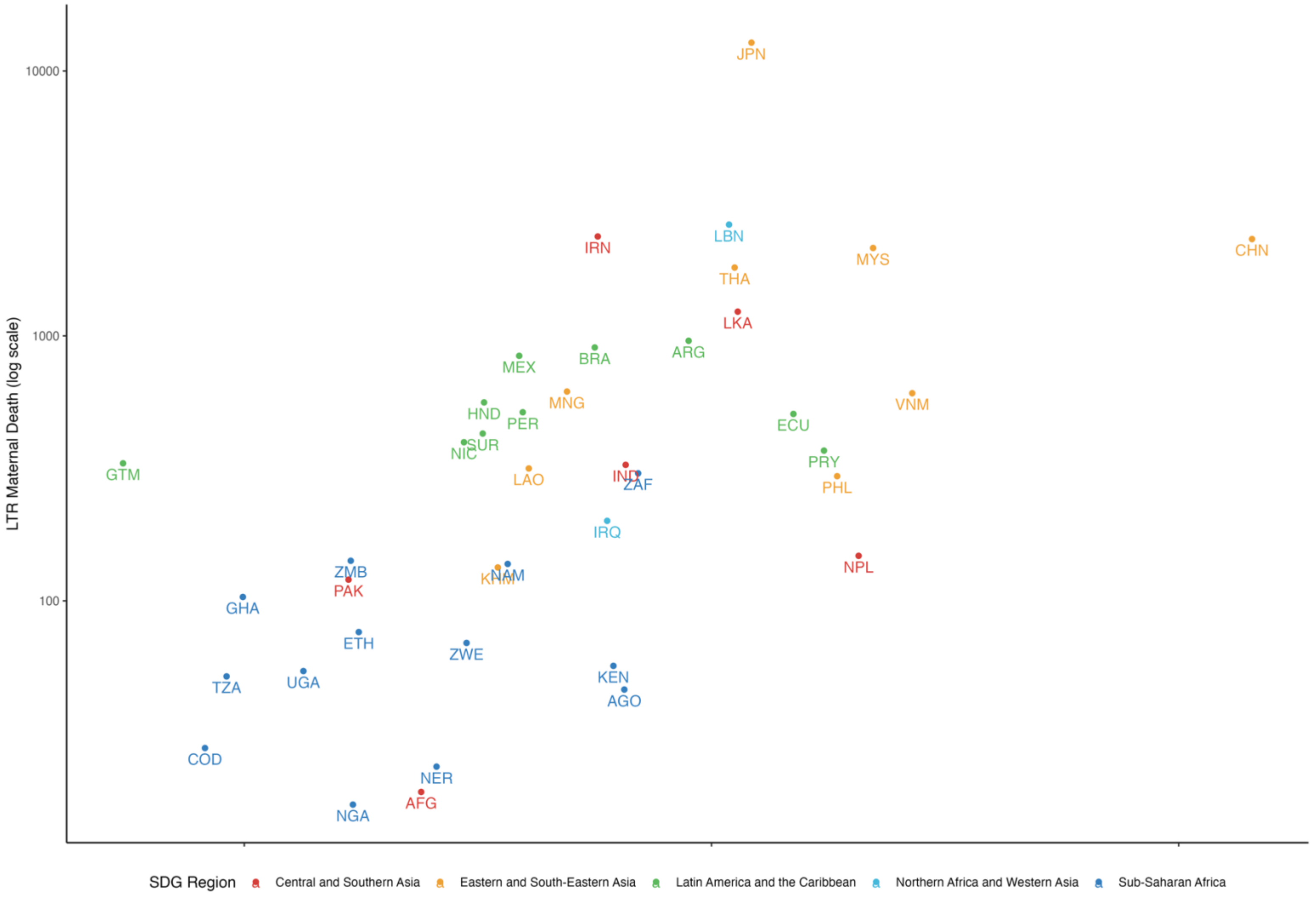

#### 5. Sensitivity Analyses

Table S4 (below) presents our calculation of the LTR-MNM where the denominator of facility-based studies has or has not been adjusted by the institutional delivery rate prior to meta-analysis pooled MNM ratio estimation (see Table S3). We also show the percentage difference in the resulting LTR-MNM estimate with and without facility-based denominator adjustment. The effect of our adjustment is large and heterogeneous by region. It has a much greater effect in regions such as sub-Saharan Africa where institutional delivery rates are low and population-based surveillance data are scarce.

This results in a reduction of the estimated level of obstetric risk for many studies conducted in low resource settings, and consequently, results in a lower estimate of the LTR-MNM than would be the case if this adjustment was not applied.

However, despite the substantial effect of our adjustment, this is preferable to using the unadjusted MNM ratio in our meta-analyses and LTR-MNM calculation. For countries with low institutional delivery rate, the number of births recorded in a tertiary facility is not representative of the number of births in the population, and therefore results in an overestimation of the MNM ratio and hence also the LTR-MNM. Since we can only adjust for the institutional delivery rate, and not according to the level of facility (primary, secondary, tertiary), this is an imperfect adjustment, but preferable to no adjustment at all.

**Table 4 Sensitivity of lifetime risk of maternal near miss to adjustment of facility-based studies**

| | Country | Year midpoint | LTR-MNM Denom. Adjusted | LTR-MNM Denom. Unadjusted | Difference in LTR-MNM (%) |
|---|---|---|---|---|---|
| **Central and Southern Asia** | | | | | |
| | Afghanistan | 2010 | 24 | 10 | 57.2 |
| | India | 2014 | 66 | 46 | 29.2 |
| | Iran | 2014 | 57 | 54 | 4.7 |
| | Nepal | 2012 | 206 | 103 | 50.2 |
| | Pakistan | 2013 | 17 | 7 | 56.8 |
| | Sri Lanka | 2010 | 114 | 113 | 0.5 |
| Mean | — | — | — | — | 33.1 |
| **Eastern and South-Eastern Asia** | | | | | |
| | Cambodia | 2010 | 35 | 29 | 16.8 |
| | China | 2015 | 1,436 | 1,436 | 0.0 |
| | Japan | 2010 | 122 | 122 | 0.2 |
| | Laos | 2020 | 41 | 41 | 0.0 |
| | Malaysia | 2014 | 222 | 222 | 0.0 |
| | Mongolia | 2010 | 49 | 48 | 1.4 |
| | Philippines | 2010 | 186 | 114 | 38.8 |
| | Thailand | 2010 | 112 | 112 | 0.3 |
| | Vietnam | 2010 | 269 | 249 | 7.5 |
| Mean | — | — | — | — | 7.2 |
| **Latin America and the Caribbean** | | | | | |
| | Argentina | 2012 | 89 | 89 | 0.6 |
| | Brazil | 2011 | 56 | 56 | 1.0 |
| | Ecuador | 2010 | 150 | 130 | 13.1 |
| | Guatemala | 2016 | 6 | 6 | 0.0 |
| | Honduras | 2014 | 33 | 24 | 28.0 |
| | Mexico | 2010 | 39 | 37 | 4.4 |
| | Nicaragua | 2010 | 30 | 21 | 29.0 |
| | Paraguay | 2010 | 174 | 166 | 4.5 |
| | Peru | 2010 | 40 | 35 | 11.4 |
| | Suriname | 2018 | 32 | 30 | 7.1 |
| Mean | — | — | — | — | 9.9 |
| **Northern Africa and Western Asia** | | | | | |
| | Iraq | 2010 | 60 | 46 | 23.5 |
| | Lebanon | 2010 | 109 | 105 | 3.3 |
| Mean | — | — | — | — | 13.4 |
| **Sub-Saharan Africa** | | | | | |
| | Angola | 2010 | 65 | 30 | 54.4 |
| | Democratic Republic of Congo | 2013 | 8 | 7 | 15.4 |
| | Ethiopia | 2018 | 18 | 6 | 68.4 |
| | Ghana | 2016 | 10 | 8 | 21.3 |
| | Kenya | 2015 | 62 | 38 | 39.0 |

| | Country | Year midpoint | LTR-MNM Denom. Adjusted | LTR-MNM Denom. Unadjusted | Difference in LTR-MNM (%) |
|---|---|---|---|---|---|
| | Namibia | 2018 | 37 | 36 | 1.6 |
| | Niger | 2010 | 26 | 8 | 70.2 |
| | Nigeria | 2014 | 17 | 6 | 64.2 |
| | South Africa | 2014 | 70 | 70 | 0.0 |
| | Tanzania | 2012 | 9 | 6 | 37.4 |
| | Uganda | 2012 | 13 | 9 | 35.5 |
| | Zambia | 2016 | 17 | 17 | 0.0 |
| | Zimbabwe | 2016 | 30 | 30 | 0.0 |
| Mean | — | — | — | — | 31.3 |

#### 6. Uncertainty

We estimated uncertainty in the LTR-MNM that derive from underlying uncertainty in the estimate of the MNM ratio only. This is to understand the contribution of uncertainty in the prevalence of MNM to the resulting estimates of the LTR-MNM and does not account for additional potential uncertainty that derives from the fertility and mortality estimates. World Population Prospects do not publish the uncertainty in their lifetable estimates.

Table S5 presents the MNM ratio and LTR-MNM with their corresponding 95% confidence intervals in parentheses. For countries where there is a large degree of variability in the MNM ratio estimates across studies, this corresponds to substantial uncertainty in the pooled MNM ratio estimate, and hence also in the LTR-MNM estimate. This emphasises the heterogeneity in MNM prevalence study design and measurement.

**Table 5 Uncertainty in the estimates of the lifetime risk of maternal near miss**

| Country | Year midpoint | No. of studies in MNM ratio meta-analysis | MNM ratio (95% CI) | LTR-MNM 1 in N (95% CI) |
|---|---|---|---|---|
| **Central and Southern Asia** | | | | |
| Afghanistan | 2010 | 1 | 7.1 (6.5, 7.9) | 24 (26, 22) |
| India | 2014 | 4 | 6.7 (2.5, 18.3) | 66 (179, 24) |
| Iran | 2014 | 3 | 8.7 (2.3, 32.7) | 57 (215, 15) |
| Nepal | 2012 | 2 | 2.1 (1.8, 2.4) | 207 (235, 181) |
| Pakistan | 2013 | 2 | 14.8 (0.7, 336.2) | 17 (381, 1) |
| Sri Lanka | 2010 | 1 | 4.0 (3.2, 5.1) | 114 (143, 91) |
| **Eastern and South-Eastern Asia** | | | | |
| Cambodia | 2010 | 1 | 10.6 (8.2, 13.7) | 35 (45, 27) |
| China | 2015 | 6 | 0.4 (0.1, 3.2) | 1436 (10049, 191) |
| Japan | 2010 | 1 | 5.9 (3.9, 9.1) | 122 (186, 79) |
| Laos | 2020 | 1 | 9.8 (5.4, 17.6) | 41 (73, 23) |
| Malaysia | 2014 | 1 | 2.2 (1.6, 2.9) | 222 (295, 167) |
| Mongolia | 2010 | 1 | 8.2 (6.4, 10.6) | 49 (63, 38) |
| Philippines | 2010 | 1 | 1.7 (1.2, 2.4) | 186 (267, 129) |
| Thailand | 2010 | 1 | 5.7 (4.3, 7.5) | 112 (147, 85) |
| Vietnam | 2010 | 1 | 2.0 (1.4, 2.8) | 269 (378, 192) |
| **Latin America and the Caribbean** | | | | |
| Argentina | 2012 | 3 | 4.8 (3.9, 6.0) | 89 (111, 72) |
| Brazil | 2011 | 4 | 10.0 (3.4, 29.4) | 56 (166, 19) |
| Ecuador | 2010 | 1 | 2.6 (1.8, 3.7) | 150 (213, 105) |
| Guatemala | 2016 | 1 | 61.9 (58.7, 65.4) | 6 (6, 5) |
| Honduras | 2014 | 1 | 11.8 (6.3, 21.8) | 33 (60, 18) |
| Mexico | 2010 | 1 | 11.1 (9.5, 13.0) | 39 (45, 33) |
| Nicaragua | 2010 | 1 | 13.2 (11.0, 15.7) | 30 (35, 25) |
| Paraguay | 2010 | 1 | 2.1 (1.1, 4.2) | 174 (350, 87) |
| Peru | 2010 | 1 | 10.0 (8.6, 11.6) | 39 (46, 34) |
| Suriname | 2018 | 3 | 12.9 (6.4, 25.9) | 32 (65, 16) |
| **Northern Africa and Western Asia** | | | | |
| Iraq | 2010 | 1 | 3.9 (3.2, 4.6) | 60 (71, 50) |
| Lebanon | 2010 | 1 | 4.3 (2.7, 6.9) | 109 (173, 69) |
| **Sub-Saharan Africa** | | | | |
| Angola | 2010 | 1 | 2.6 (2.0, 3.4) | 65 (85, 50) |
| Democratic Republic of Congo | 2013 | 2 | 19.7 (4.4, 88.4) | 8 (37, 2) |
| Ethiopia | 2018 | 8 | 13.5 (4.7, 38.5) | 18 (50, 6) |
| Ghana | 2016 | 1 | 26.9 (24.0, 30.1) | 10 (11, 9) |
| Kenya | 2015 | 3 | 4.5 (0.3, 56.1) | 62 (784, 5) |
| Namibia | 2018 | 2 | 8.2 (7.4, 9.1) | 37 (41, 33) |
| Niger | 2010 | 1 | 5.5 (4.7, 6.3) | 26 (30, 22) |
| Nigeria | 2014 | 3 | 11.3 (3.4, 37.2) | 17 (56, 5) |
| South Africa | 2014 | 3 | 6.1 (4.2, 8.9) | 70 (101, 48) |
| Tanzania | 2012 | 1 | 22.3 (20.4, 24.4) | 9 (10, 8) |

| Country | Year midpoint | No. of studies in MNM ratio meta-analysis | MNM ratio (95% CI) | LTR-MNM 1 in N (95% CI) |
|---|---|---|---|---|
| Uganda | 2012 | 2 | 13.6 (4.4, 41.9) | 13 (41, 4) |
| Zambia | 2016 | 1 | 13.0 (11.2, 15.1) | 17 (20, 15) |
| Zimbabwe | 2016 | 1 | 9.3 (7.7, 11.2) | 30 (36, 25) |
